# Impact of Mass Distribution of Long-Lasting Insecticide Nets on Malaria Prevention in Lindi Region, Tanzania: A quasi experimental study

**DOI:** 10.1101/2024.12.09.24318122

**Authors:** Epafra Luka Mwanja, Scola Anilozi Mwalyanzi, Seif S. Khalfan, Ezekia Jasson Ambikile, Mgole Eliud Mkama, Dotto Daniel Kisendi

## Abstract

**Background:** Malaria is a major public health issue with high rates of morbidity and mortality in the United Republic of Tanzania. To ensure that all households remain protected, national malaria control programs and partners in 2022 implemented free mass distribution of long-lasting insecticides nets to population at risk and high prevalent regions with malaria including Lindi region. The present study aimed to assess the effectiveness of free long-lasting insecticide-treated nets in reducing malaria burden among the population.

**Methods:** we conducted a quasi-experimental study from September 2021 to August 2022 and September 2022 to August 2023. The data were collected on District Health Information System version two in Lindi region and analysed with T-tests to compare the malaria positive rate before (September 2021 to August 2022) and after the distribution of long-lasting insecticides nets (September 2022 to August 2023). Malaria positive rate in the general population who attended outpatient department was reduced by 7.6% after distribution of long-lasting insecticides nets. A comparison of malaria cases recorded between September 2021 and August 2023 in the different area showed disparities. Before long lasting insecticides nets implementation, the malaria positive rate in all area combined was 20.6%. Whereas malaria positive rate in all area combined was 13% after implementation. The mean difference is 6.60, with a standard deviation of 2.47. The t-value is 6.55, with 5 degrees of freedom. The significance (2-tailed) value is <0.001, indicating a statistically significant difference between the malaria positive rate before and after long lasting insecticides nets implementation.

**Conclusion:** the long-lasting insectides nets distribution campaign synergy with other preventive interventions has had a significant impact on reducing the malaria positive rate in the population.

## Introduction

Malaria remains a critical public health challenge in Tanzania, characterized by high morbidity and mortality rates, similar to many sub-Saharan African nations. The World Health Organization (WHO) reported an estimated 10.8 million malaria cases in Tanzania in 2019, representing 5% of global cases (1). The disease is transmitted by infected mosquitoes, and long-lasting insecticidal nets (LLINs) are recognized as one of the most effective preventive measure (2). LLINs not only repel but also kill mosquitoes upon contact, significantly reducing the risk of malaria transmission (3). In Tanzania, mass distribution of LLINs has been a key strategy to enhance coverage within communities (4).

Initially, LLIN programs focused on the most vulnerable groups: children under 5 years and pregnant women. This combined approach of social marketing and distribution led to a notable increase in net usage, with 64% of children under 5 and 66% of pregnant women sleeping under LLINs by 2022 (5). Over five years, this initiative contributed to a decline in malaria incidence from 162 per 1,000 population in 2015 to 106 in 2020 (6). The National Malaria Control Programme (NMCP) and its partners achieved universal LLIN coverage through extensive door-to-door distribution campaigns from 2010 to 2011 (7), reaching 23 out of 26 mainland regions by 2015 and targeting 50 councils across 12 regions in 2020. The objective was to ensure that all individuals, regardless of age or sex, had access to LLINs, aiming for at least 80% protection of the at-risk population. The use of LLINs is intended to interrupt malaria transmission by eliminating the human reservoir of the parasite, thereby reducing community-level risk (5). In 2010-11, to sustain universal LLINs coverage, the NMCP initiated a mixed model of LLINs distribution. Alongside with continuous distribution mechanisms using antenatal care, child welfare clinics, schools, shops and workplace programs and mass distribution of free LLINs every three years since 2012 (7).

In 2022, to further protect at-risk populations in high-prevalence regions, including Lindi (12% malaria prevalence), Mtwara (15%), Kigoma (24%), Geita (17%), Kagera (15%), and Ruvuma (12%), the NMCP implemented a free mass distribution campaign (8). On September 26, 2022, over 710,370 LLINs were distributed in Lindi alone, aiming for a coverage rate of at least 90% among the general population. Despite these efforts, malaria morbidity in the Lindi region remains high. This study aims to assess the malaria positivity rate before and after the mass distribution of LLINs in Lindi, addressing the existing evidence gap in this area.

## Methods

### Study design and setting

We conducted a quasi-experimental study from September 2021 to August 2022 and September 2022 to August 2023 in Lindi Region. It is one of 28 regions that make up the United Republic of Tanzania, located in the Southern zone of Tanzania, divided into 6 districts council. All health facilities within, Kilwa, Liwale, Lindi municipal, Mtama, Ruangwa and Nachingwea districts council of Lindi region were included in this study. Because individuals who received LLINs and who did not receive LLINs seek malaria prevention and treatment service in these health facilities. In this study all health facilities in Mtama, Ruangwa and Nachingwea districts council of Lindi region that participated in mass distribution of LLINs were assigned into intervention group, and all health facilities in Lindi municipal, Kilwa districts and Liwale districts of Lindi region that did not receive the LLINs were assigned into control group. The year 2021 was used as a baseline.

### Study population, variables and study size

Exhaustive sampling was used in this study. It included the general population in both urban and rural area who attended at outpatient department to seek malaria prevention and treatment service in all health facilities at those selected area, Kilwa, Liwale, Lindi municipal, Mtama, Ruangwa and Nachingwea districts of Lindi region. The malaria positive rate among all OPD cases across all health facilities per year before and after LLINs implementation, the number of confirmed malaria cases among OPD cases across all health facilities per year, were the outcome variable for this study. A total of 2,153,208 participants who attended at OPD for malaria prevention and treatment service in all health facilities were recruited between September 2021 to August 2022 and September 2022 to August 2023 in Lindi Region.

### Case standard definition

#### Confirmed malaria case

A confirmed malaria case is defined as a person who attended an outpatient department at any health facility within Lindi region and had a positive rapid diagnostic test (RDT) or a positive microscopy result for malaria parasites.

#### Malaria positive rate

The malaria positive rate is defined as the proportion of confirmed malaria cases among the total population tested for malaria at outpatient department in health facilities within Lindi region on their respective selected area/districts.

Formula

**Malaria Positive Rate = (Number of Confirmed Malaria Cases / Total Number of Person Tested for Malaria at Outpatient Department) x 100**.

Where:

Number of Confirmed Malaria Cases: The number of individuals who had a positive rapid diagnostic test (RDT) or a positive microscopy result for malaria parasites.

Total Number of Persons Tested for Malaria at Outpatient Department: The total number of individuals who underwent either an RDT or microscopy test for malaria at the outpatients department in any health facilities within Lindi region. The malaria positive rate were expressed as a percentage (%), indicating the proportion of tested individuals who were confirmed to have malaria.

### Data collection, bias, quantitative variables

The data analyzed in this study were taken from DHIS2, which is the national health data management software that Lindi Region has been using for several years ago. Poor data completeness and coding errors on the DHIS2 may cause bias and influence the results of the study. However, DHIS2 standards state that when data completeness is 80% or more, analyzes can be done. The collected data are number of OPD attendance, number of confirmed malaria cases in OPD (confirmed by a rapid diagnostic test or thick film smear) and malaria positive rate in each area before and after LLINs implementation. Also, number of confirmed malaria cases, using the rapid diagnostic testing (RDT) or the microscopy testing or both.

### Statistical methods

Data were exported to Microsoft Excel and analyzed using SPSS for figure and table development, as well as result interpretation. Malaria positivity rates were calculated for two periods: before the mass distribution of LLINs (September 2021 to August 2022) and after (September 2022 to August 2023). These rates were compared across the entire Lindi region and by individual districts. Specifically, districts participating in LLIN distribution (Mtama, Ruangwa, and Nachingwea) were compared to those in Lindi municipal, Kilwa, and Liwale that did not receive LLINs. T-tests were employed to assess differences in malaria positivity rates among the populations. A paired sample T-test in SPSS was used to calculate p-values, with significance set at p < 0.05.

## Results

### Demographic characteristics of enrolled participants

A total of 2,153,208 participants who attended at OPD for health service were recruited between September 2021 to August 2022 (before ITN implementation) and September 2022 to August 2023 (after ITN implementation) in Lindi Region. In regards to sex 1,202,923 participants were female and 950,285 participants were male. In term of age group 992,765 participants were under 5 years old and 1,160,443 participants were above 5years (Table 2).A total of 1,096217 participants were enrolled between September 2021 to August 2022 (before ITN implementation), where total of 1,056,991 were enrolled between September 2022 to August 2023 (after ITN implementation) (Table 1).

**Table 1:**
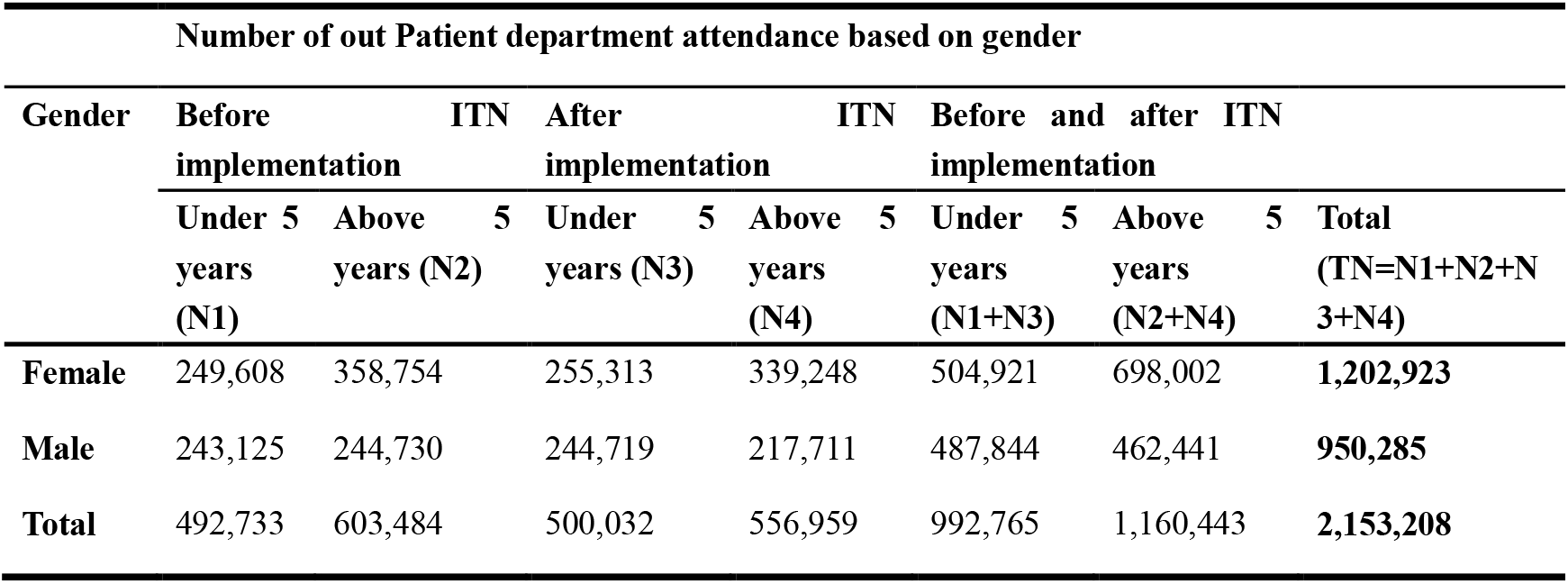
Demographic characteristics of enrolled participants.

**Table 2:**
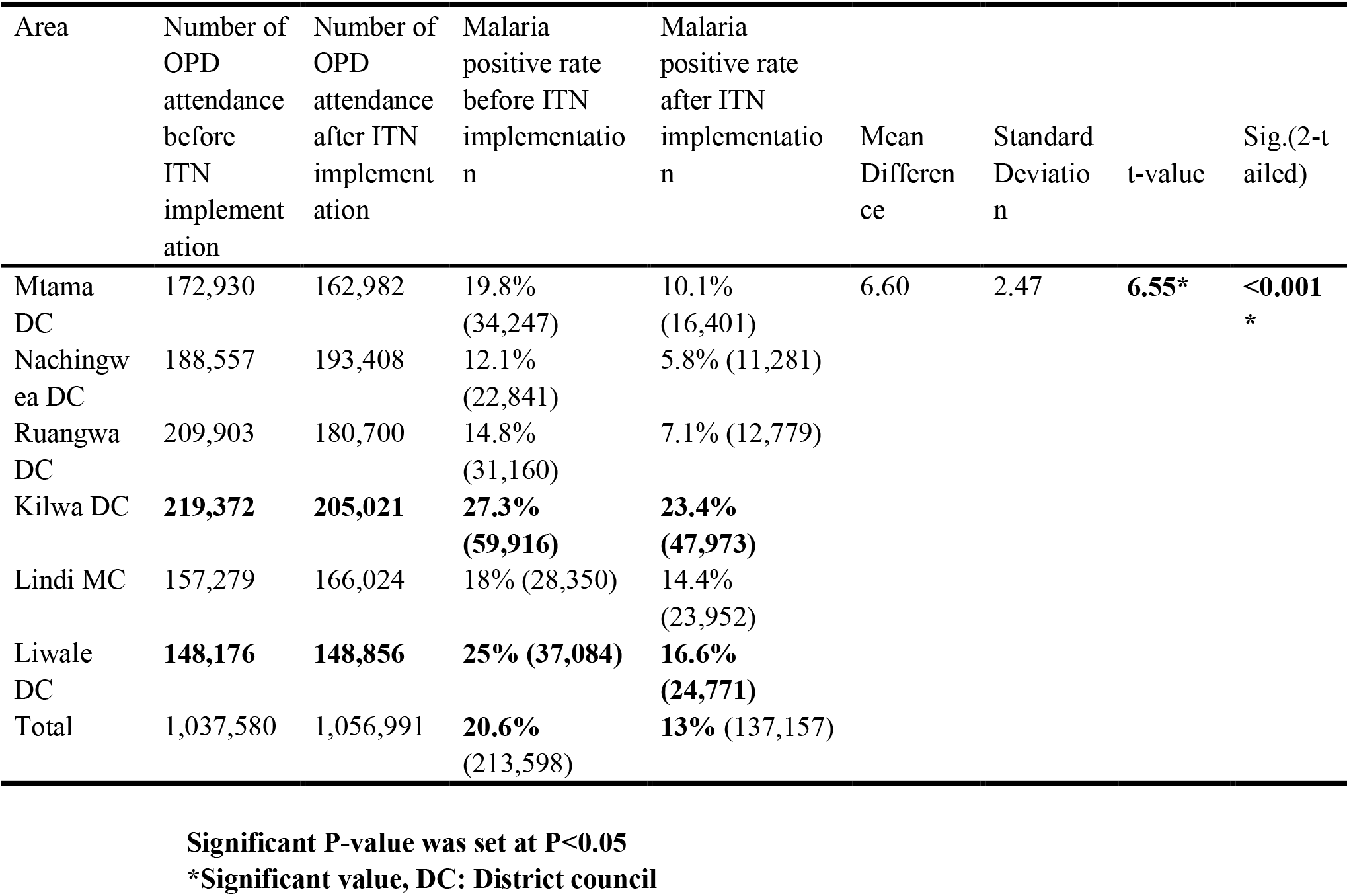
Number of OPD attendance before and after LLINs implementation, Malaria positive rates before and after implementation across all areas, and T-test.

| Area | Number of OPD attendance before ITN implementation | Number of OPD attendance after ITN implementation | Malaria positive rate before ITN implementation | Malaria positive rate after ITN implementation | Mean Difference | Standard Deviation | t-value | Sig.(2-tailed) |
| --- | --- | --- | --- | --- | --- | --- | --- | --- |
| Mtama DC | 172,930 | 162,982 | 19.8% (34,247) | 10.1% (16,401) | 6.60 | 2.47 | **6.55\*** | **<0.001\*** |
| Nachingwea DC | 188,557 | 193,408 | 12.1% (22,841) | 5.8% (11,281) |  |  |  |  |
| Ruangwa DC | 209,903 | 180,700 | 14.8% (31,160) | 7.1% (12,779) |  |  |  |  |
| Kilwa DC | **219,372** | **205,021** | **27.3% (59,916)** | **23.4% (47,973)** |  |  |  |  |
| Lindi MC | 157,279 | 166,024 | 18% (28,350) | 14.4% (23,952) |  |  |  |  |
| Liwale DC | **148,176** | **148,856** | **25% (37,084)** | **16.6% (24,771)** |  |  |  |  |
| Total | 1,037,580 | 1,056,991 | **20.6% (213,598)** | **13% (137,157)** |  |  |  |  |

**Significant P-value was set at P<0.05**
**\*Significant value, DC: District council**

### Number of OPD attendance before and after LLINs implementation, Malaria positive rates before and after implementation

According to Table 2, the results show outpatient department (OPD) attendance from September 2021 to August 2022, prior to LLIN implementation varied between 148,176 and 219,372, while confirmed malaria cases ranged from 22,841 to 59,916. Malaria positivity rates across areas ranged from 12.1% to 27.3%. Kilwa District Council had the highest OPD attendance and confirmed cases, with a positivity rate of 27.3%. Overall, the combined malaria positivity rate across all areas was 20.6%.

Data on outpatient department (OPD) attendance from September 2022 to August 2023, following the implementation of long-lasting insecticidal nets (LLINs) ranged from 148,856 to 205,021, while confirmed malaria cases varied from 11,281 to 47,973. The malaria positivity rates across areas ranged from 5.8% to 23.4%, with a total OPD attendance of 1,056,991 and 137,157 malaria cases. Kilwa DC had the highest OPD attendance and confirmed malaria cases (23.4%), while Nachingwea DC reported the lowest positivity rate (5.8%). The combined malaria positivity rate across all areas was 13% after LLINs implementation. Generally, OPD attendance decreased after LLIN implementation. The trends across Mtama DC, Nachingwea DC, Ruangwa DC, Kilwa DC, Lindi MC, and Liwale DC indicate a reduction in malaria positivity rates following LLIN implementation. This suggests a positive impact of LLINs on reducing malaria cases in all areas. Figure 1 illustrates these trends, highlighting the overall decline in malaria positive rates after LLINs were introduced. The implementation of LLINs has effectively contributed to lower OPD attendance and reduced malaria positivity rates. Regarding compare means paired sample T-test (Table 2) comparing the malaria positive rate before and after the implementation of

**Figure 1:**
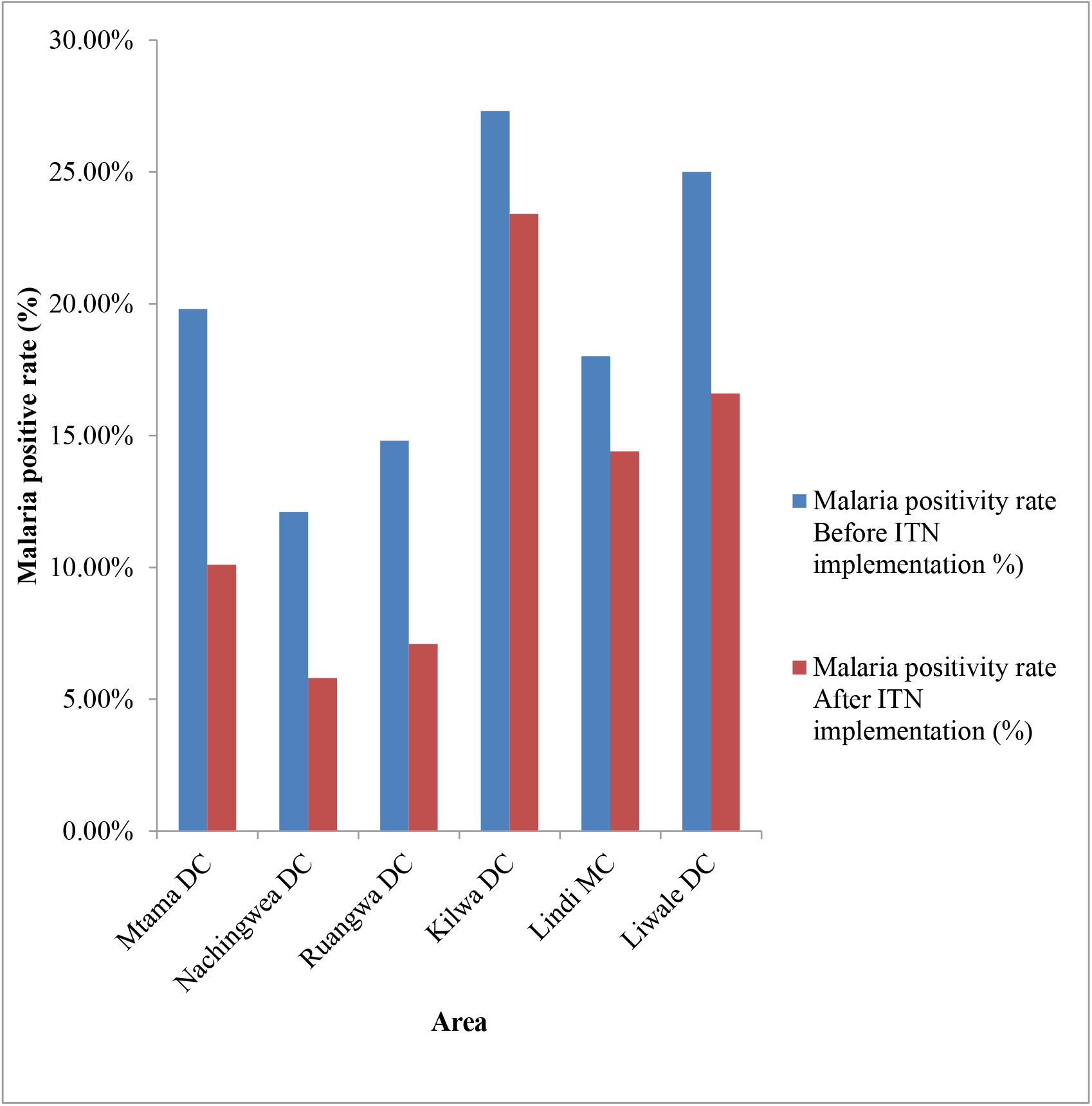
Malaria positive rates before and after implementation across all areas. NB; DC-district council, MC-Municipal council

LLINs, the paired differences show a mean difference of 6.60 in malaria positivity rates before and after LLIN implementation, with a standard deviation of 2.47 and a T-value of 6.55. The significant p-value of <0.001 indicates LLINs effectively reduced malaria burdens, highlighting the need for further interventions.

### Comparison between experimental group (Area received LLINs) and control group (Area that did not receive LLINs)

Table 3 compares the experimental group (areas receiving LLINs) with the control group (areas not receiving LLINs) regarding malaria positive rates before and after intervention. The experimental group includes Mtama DC, Nachingwea DC, and Ruangwa DC, while the control group consists of Kilwa DC, Lindi MC, and Liwale DC. Before the LLINs intervention, the malaria positive rate was 15.4% in the experimental group and 23.9% in the control group. After the intervention, the rates decreased to 7.5% in the experimental group and 18.6% in the control group. The LLINs intervention effectively reduced malaria rates in both groups, with the experimental group consistently showing lower rates.

**Table 3:**
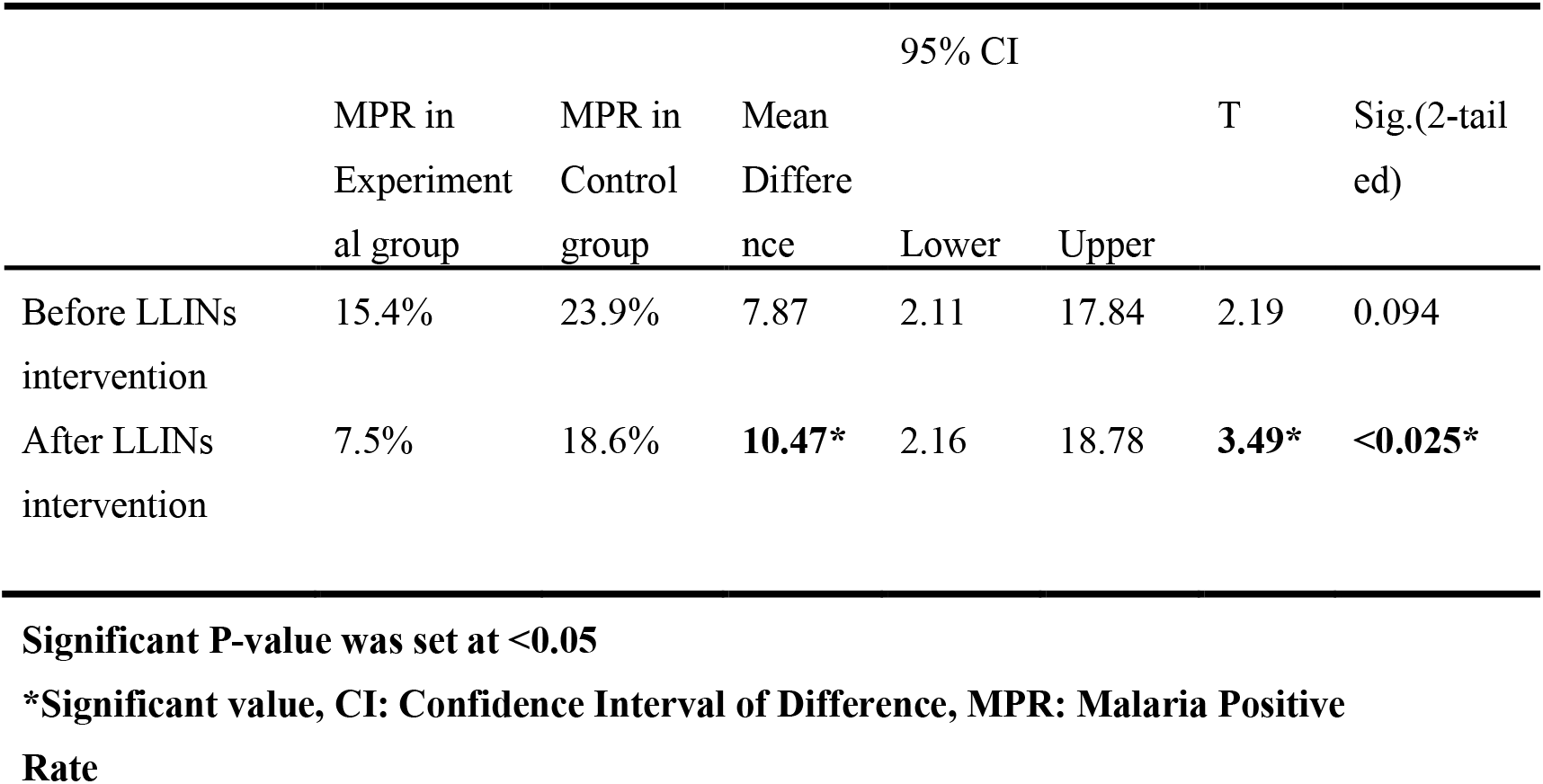
Malaria positive rate in experimental and control group before LLINs interventions, and respective independent sample T test.

Additionally, an independent samples t-test revealed a t-statistic of 3.49 and a p-value of 0.025, indicating a statistically significant difference between the groups after LLIN implementation. The mean difference was 10.47, with a 95% confidence interval ranging from 2.16 to 18.78.

### Malaria Positive Rate

The Figure 2 compares the malaria positive rate before and after mass distribution of LLINs in Lindi Region. Before the LLINs intervention, the malaria positive rate ranges from 18.9% to 50.9%, where the month with highest malaria positive rate June 2022 (50.9%). After the LLINs intervention, the malaria positive rate ranges from 18.1% to 31.4%, where the month with highest malaria positive rate June 2022 (31.4%). The LLINs intervention resulted in a decrease in the malaria positive rate. After LLINs interventions results show lower malaria positive rate compared to before intervention. The LLINs intervention was effective in reducing malaria positive rate in the study area.

**Figure 2:**
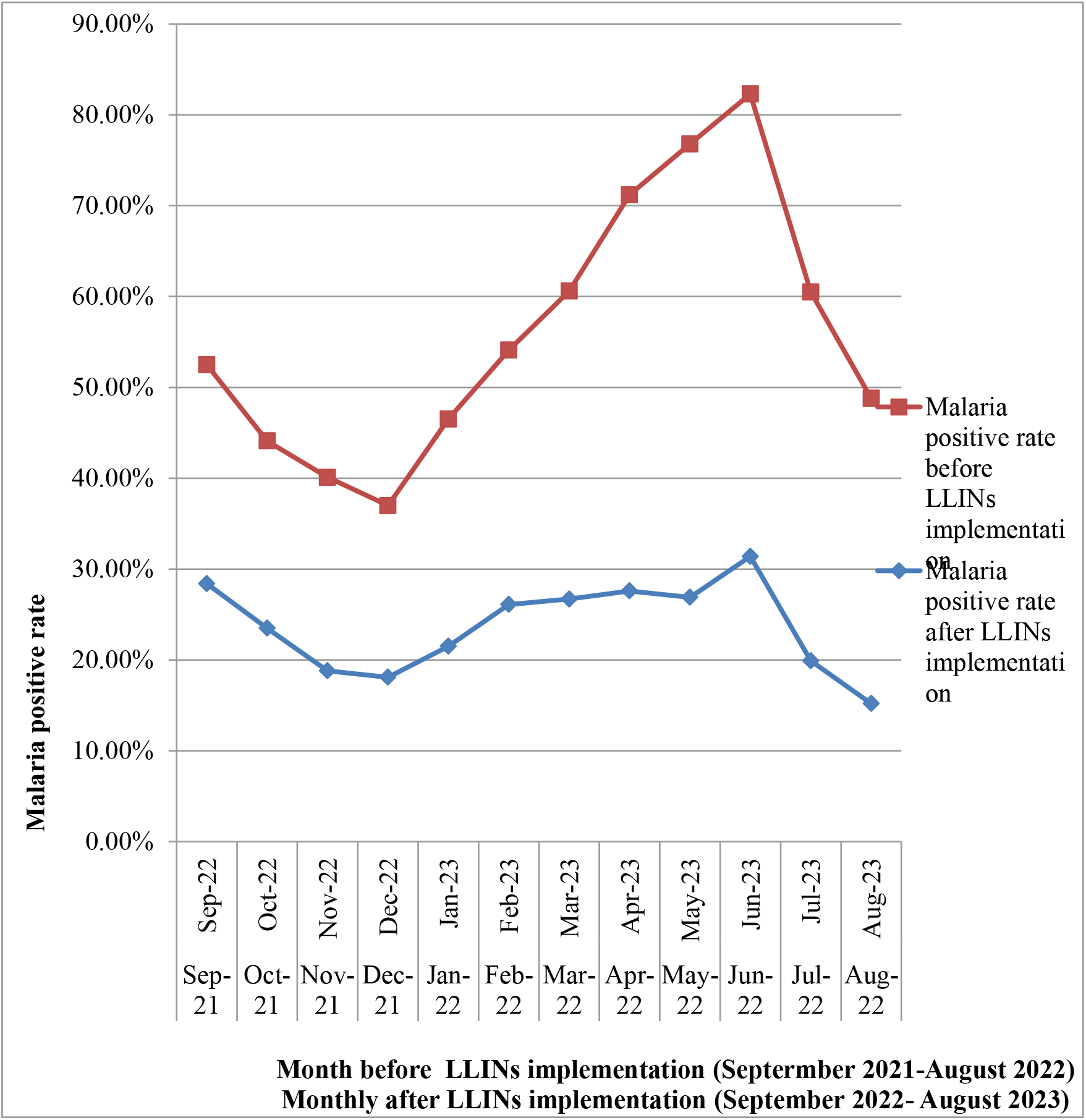
Trends of Malaria positive rate before and after mass distribution of Long-Lasting Insecticides Nets between September 2021-August 2022 and September 2022-August 2023 in Lindi region.

## Discussion

In 2022, the Tanzania Ministry of Health launched a free distribution campaign for long-lasting insecticidal nets (LLINs) in malaria-prone regions, including Lindi. In September 2022, 710,370 LLINs were distributed across the most affected districts. This mass distribution led to a significant decrease in malaria cases, with the malaria positive rate in the general population dropping by 7.6% from September 2022 to August 2023. Before LLINs implementation, malaria positive rates ranged from 12.1% to 27.3%, with an overall rate of 20.6%. After the intervention, rates decreased to between 5.8% and 23.4%, resulting in a combined rate of 13%. The observed decline in malaria cases can be attributed to several factors. Following the campaign, the population was aware of the efficacy and proper use of LLINs due to prior educational sessions. Correct utilization was noted, and the insecticides used for impregnation remained effective.

However, there was a concerning increase in malaria positive rates from 26.1% to 31.4% between February and June 2023. Several studies indicate that the effectiveness of LLINs varies post-distribution. A study in Tanzania found that after three years, less than 17% of LLINs were still usable, with functional survival times of 1.6 years for LLINs containing pyrethroid and piperonyl-butoxide (PBO), and 1.9 years for standard LLINs (9). Nevertheless, PBO LLINs offered more protection than standard ones, regardless of condition (10). Another studies conducted in Tanzania, India, and Côte d’Ivoire assessed the durability of pyrethroid-PBO LLINs and found that their field durability met the World Health Organization’s (WHO) criteria for being categorized as “long-lasting”(11). These findings highlight the importance of monitoring LLINs’ durability and considering the replenishment intervals for PBO nets to ensure their continued effectiveness in malaria control programs.

Several studies demonstrate the effectiveness of LLINs in preventing malaria. LLINs reduce vector density and disease transmission by preventing contact between mosquitoes and individuals (12). However, the effectiveness of LLINs can decrease over time, with reduced mortality rates observed after three years of usage(13). It is recommended to re-dip LLINs with insecticide doses to maintain their effectiveness. While ownership of LLINs is high, usage levels can be moderately low, indicating the need to address barriers to consistent utilization (14). The durability of LLINs is an important factor, and there is a need for more durable nets that last longer than the current 2-3-year rating. The comparative analysis of the incidence rates between the two periods (before and after mass distribution of LLINs) shows that there is statistically significant difference. In other words, the LLINs distribution campaign did really reverse the epidemiological situation. This is linked to the fact that prevention of malaria in our environment didn’t tends to focus only on the use of LLINs but also other malaria preventive methods as indoor residual spraying, environmental management, early case treatment and personal protection. However, there are challenges associated with distribution, increasing pyrethroid-resistant mosquito population, access, shorter lifespan of LLINs and use of LLINs at the household level that should be considered during allocation of scarce financial resources for populations. This analysis shows that high malaria positive rate is concentrated in kilwa districts of Lindi region; however the free LLINS was not distributed in that area. If funding gaps continue, targeted selection of high burden, more populated regions and districts should be prioritized.

Malaria elimination in Tanzania is a public health problem and requires a comprehensive approach. The World Health Organization (WHO) defines elimination as the absence of recent indigenous cases in an area (15). To achieve malaria elimination, this study recommends strengthening malaria surveillance and reporting, reactive case detection, and reactive drug administration (16). The 1,7-malaria Reactive Community-based Testing and Response (1,7-mRCTR) approach, adapted from the 1-3-7 strategy, should be implemented. The adapted 1,7-mRCTR strategy calls for reporting confirmed malaria cases at health facilities within one day and conducting community-wide testing in selected villages within seven days to slow transmission during the same phase of the Plasmodium life cycle (17). Where 1-3-7 approach involves reporting cases within one day, investigating them within three days, and responding to foci within seven days. This strategy has been implemented in several countries, including China(18), Hainan Province in China (19), and Thailand (20,21). The strategy has shown success in reducing malaria incidence and accelerating progress towards elimination. Key components include active case detection, vector surveillance, and community-based health workers (22). Support through training, capacity building, and knowledge dissemination is essential for successful implementation. A comprehensive national effort, with backing from various government ministries, is necessary for operationalizing malaria elimination plans. Additionally, expanding coverage of malaria prevention interventions, particularly increasing the distribution and utilization of long-lasting insecticidal nets (LLINs), is vital. Strengthening community engagement and behavior change communication will promote early care-seeking, proper LLIN use, and adherence to malaria treatment.

The study had some limitations. First, selection of study sites was based on data availability and given the small number of health facilities in the selected districts, the study may not have had sufficient statistical power to detect differences between health facilities received LLINs and that not received, as evidenced by the wide confidence intervals around effect estimates. Second, though the DHIS2 database enabled us to capture a comprehensive set of delivery information, poor data completeness and coding errors on the DHIS2 can lead to bias and influence the results. Third, geographic information on the residence of patients attending out patient’s department at health facilities was not collected due to frequent absence of this variable. Thus, the health organization units where the patients attended for malaria investigation may not accurately represent their health facilities of residence. Lastly, health facilities were selected non-randomly and though quasi-experimental study designs were used to control for confounding, unmeasured confounding may have biased results. Thus, estimates should be interpreted with caution.

In conclusion, the results of this study unequivocally demonstrate that the mass distribution of LLINs has significantly reduced the percentage of malaria cases in Lindi Region. This intervention has not only contributed to a decline in malaria transmission but has also led to other positive outcomes, such as a reduction in severe malaria cases and hospital admissions. The success of this intervention underscores the importance of sustained efforts in malaria prevention and control, with LLINs playing a pivotal role in achieving these goals. Moving forward, it is imperative to continue supporting and promoting the use of LLINs as part of comprehensive malaria control strategies, ensuring that these life-saving interventions reach those who need them the most.

## Data Availability

All data produced in the present study are available upon reasonable request to the authors

## Declarations

## Acknowledgements

Not applicable

## Ethical Approval to participate

Not applicable

## Consent for publication

Not applicable

## Availability of data and material

Data generated or analyzed during this study are included in this published article. All sources identified in the study are publicly available and are cited in the manuscript, meeting the requirement of data being available in the manuscript itself.

## Author’s contribution

Conceptualization and draft manuscript E.L.M.; writing—manuscript and editing, E.L.M., S.A.M., S.S.K., E.J.A.,M.E.M.,D.D.K.; All authors have read and agreed to the published version of the manuscript. E.L.M is the guarantor.

## Patients and public involvement

Patients and the public were not involved in any way

## ABBREVIATIONs

ACT: Artemisinin based combination therapy
BCC: Behavioral change communication
CQ: Chloroquine
CHW: Community health workers
CLWS: Children living and working in streets
DC: District council
DDT: Dichlorodiphenyltrichloroethane
DHP: Dihydroartemisininpiperaquine
DMTs: Districts management teams
GoT: Government of Tanzania
IMVC: Integrated malaria vector control
IEC: Information education and communications
IRS: Indoor residual spraying
LLINs: long lasting insecticidal nets
LSM: larval source management
MDGS: Millennium Developmental Goals
NMCP: National malaria control programs
PPP: Public private partnerships
PBO: Pyrethroid and piperonyl butoxide
RDT: Rapid diagnostics test
SMMSP: Supplementary malaria midterm strategic plan
TACTs: Triple artemisinin combination therapies

